# Evolving Epidemiology and Effect of Non-pharmaceutical Interventions on the Epidemic of Coronavirus Disease 2019 in Shenzhen, China

**DOI:** 10.1101/2020.05.09.20084202

**Authors:** Suli Huang, Zhen Zhang, Yongsheng Wu, Shujiang Mei, Yuan Li, Xu Xie, Xiaojian Liu, Xiujuan Tang, Dongfeng Kong, Xiaoliang Wu, Yu Wu, Lan Wei, Ziquan Lv, Shuyuan Yu, Ying Wen, Guohong Zhou, Tianmu Chen, Tiejian Feng, Xuan Zou

**Affiliations:** Department of Environment and Health, Shenzhen Center for Disease Control and Prevention, No.8 Longyuan Rd, Shenzhen 518055, China; Department of Public health information, Shenzhen Center for Disease Control and Prevention, No.8 Longyuan Rd, Shenzhen 518055, China; Department of Communicable disease control and prevention, Shenzhen Center for Disease Control and Prevention, No.8 Longyuan Rd, Shenzhen 518055, China; Department of Immune-Planning, Shenzhen Center for Disease Control and Prevention, No.8 Longyuan Rd, Shenzhen 518055, China; Department of School Hygiene, Shenzhen Center for Disease Control and Prevention, No.8 Longyuan Rd, Shenzhen 518055, China; Department of Molecular Epidemiology, Shenzhen Center for Disease Control and Prevention, No.8 Longyuan Rd, Shenzhen 518055, China; State Key Laboratory of Molecular Vaccinology and Molecular Diagnostics, School of Public Health, Xiamen University, No.422 Simin Nan Rd, Xiamen 361102, China; Shenzhen Center for Disease Control and Prevention, No.8 Longyuan Rd, Shenzhen 518055, China

**Keywords:** COVID-19, Epidemiology, Non-pharmaceutical Intervention, modeling

## Abstract

Previous studies have demonstrated the characteristics of patients with 2019 novel coronavirus disease (COVID-19). However, the effect of non-pharmaceutical interventions on the epidemic in Shenzhen, China remains unknown. Individual data of 417 cases were extracted from the epidemiological investigations and the National Infectious Disease Information System between January 1, 2020 and February 29, 2020. On the basis of important interventions, the epidemic was divided into four periods (January 1-15, January 16-22, January 23-February 5 and after February 6). We used a susceptible-exposed-infectious-asymptomatic-recovered model to evaluate the effect of interventions. Results suggested that about 53.7% were imported from Wuhan. The median age was 47 years and 52.8% were women. Severity risk increased with age and associated with male and co-existing disorders. The attack rate peaked in the third period and drastically decreased afterwards across sex, age groups and geographic regions. Children younger than 5 years showed a higher attack rate than those aged of 6~19. The effective reproductive number decreased from 1.44 to 0.05 after the highest level emergency response since January 23. Overall, the non-pharmaceutical interventions have effectively mitigated the COVID-19 outbreak in Shenzhen, China. These findings may facilitate the introduction of public health policies in other countries and regions.

## Introduction

During December 2019, a novel beta-coronavirus named SARS-CoV-2 appeared in Wuhan, China and rapidly spread across the whole country in less than a month. Until 29 February, 2020, about 79394 patients with the Coronavirus Disease 2019 (COVID-19) and 2838 deaths have been reported in China (World Health Organization). On 12 March 2020, the WHO declared the COVID-19 was a pandemic, because the virus quickly spread to 113 countries worldwide. Although increasing studies with different sample size have described the general characteristics of COVID-19 (Guan et al., 2020; Li et al., 2020), and a preliminary study has demonstrated the early transmission dynamics of COVID-19 in Shenzhen (Bi et al., 2020), the full spectrum of the epidemiological characteristics of the outbreak in Shenzhen across different time periods was warranted to illustrate.

Since the outbreak of COVID-19 in Wuhan, China, the emergency response for unexplained pneumonia has been initiated promptly since the end of December, 2019 by Shenzhen authorities and the Center for Disease Control and Prevention (CDCs). Subsequently, Shenzhen authorities and the Guangdong government have implemented a series of timely non-pharmaceutical interventions to prevent the spread of COVID-19 (**Fig 1**), including starting the joint defense and control mechanism of major infectious diseases, initiating the highest level of emergency response, and putting the “five 100%, ten must” policy into practice to control community transmission, etc.

**Figure 1.**
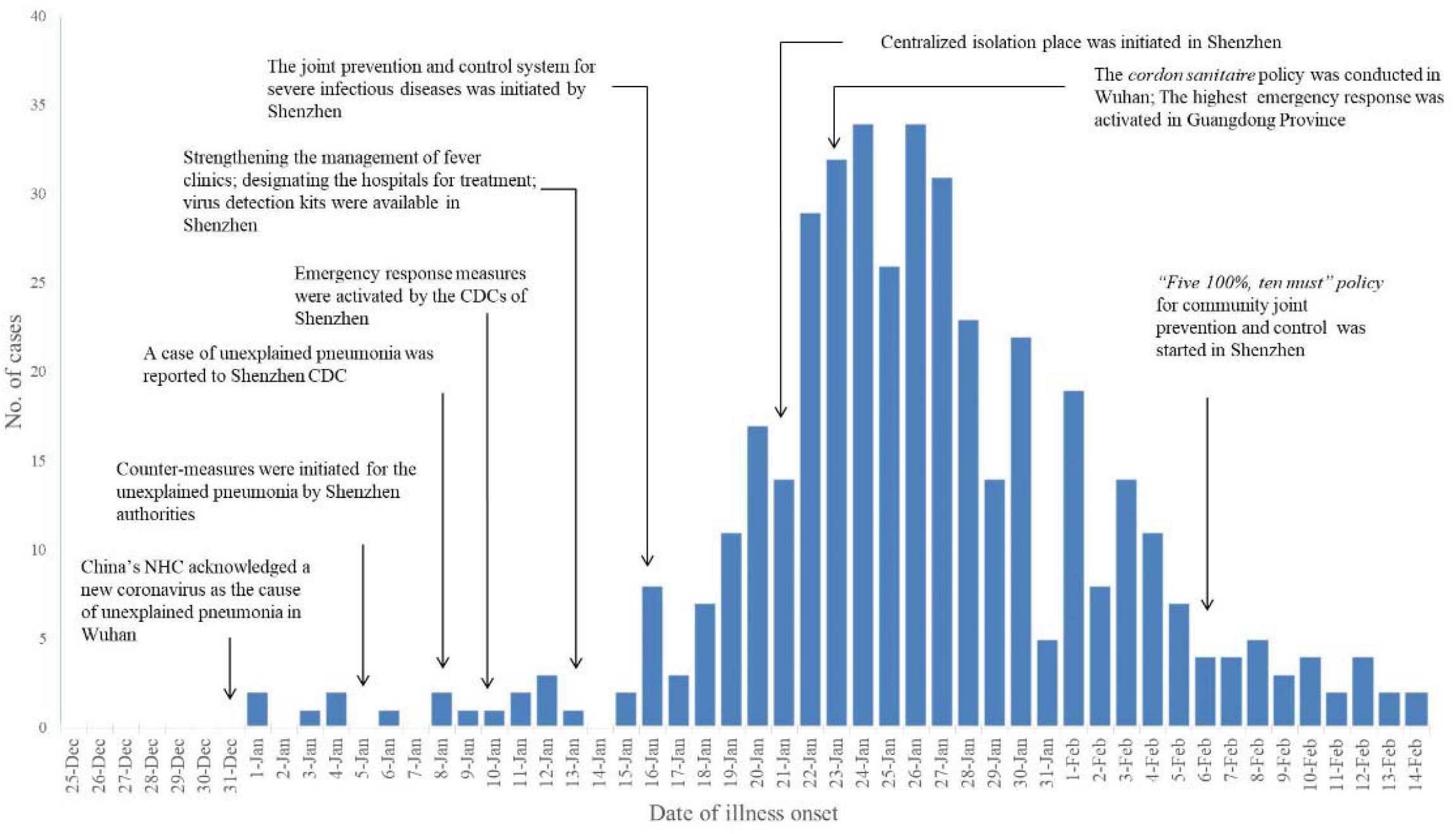
The epidemic curve of COVID-19 cases and intervention measures in Shenzhen by illness onset date.

Mathematical model was commonly used to simulate the dynamics of COVID-19 and to assess the effect of interventions (Wu et al., 2020; Zhao et al., 2020a; Prem et al., 2020). We also built a susceptible-exposed-infectious-asymptomatic-recovered (SEIAR) model to calculate the basic reproduction number (*R*_0_) and the transmissibility of COVID-19 among age groups (Chen et al., 2020; Zhao et al., 2020b). However, the modeling studies warrant more reports evaluating the effect of non-pharmaceutical interventions, which could affect the model parameters across time, such as transmission rate, duration from onset to hospitalization, etc. Increasing evidence has reported a noticeable proportion of asymptomatic patients (Hu et al., 2020; Mizumoto et al., 2020). The virus SARS-CoV-2 was assumed to be contagious both in the asymptomatic patients and during the incubation period of ascertained cases (Bai et al., 2020; Tong et al., 2020). These factors should be considered in the future modeling studies.

In this study, we described the general epidemiological characteristics of the patients with COVID-19 in different time periods in Shenzhen from January 1, 2020 to February 29, 2020. We also applied the SEIAR model to estimate the overall effect of Guangdong highest emergency response on the epidemic in Shenzhen, which might provide important information for the global combat of COVID-19.

## Results

### General characteristics

This study included a total of 417 confirmed cases, and their onset date ranged from January 1 to February 14, while the report date ranged from January 20 to February 29. The epidemic curve by the onset date, report date and important interventions is shown in **Fig 1** and **Fig S1**. Most of the cases occurred between January 18 and February 5, accounting for 85.9% of the total number. The input patients were those who lived or traveled in Hubei or from other provinces of China, and they accounted for the majority (82%) of the total patients (**Table 1**). Among the input patients, a total of 224 cases (65.5%) were from Wuhan city, but none of them was ever exposed to the Hua-Nan Seafood market, the original source of the virus. Notably, the proportion of local cases and those identified by surveillance showed an increased trend across time.

**Figure 2.**
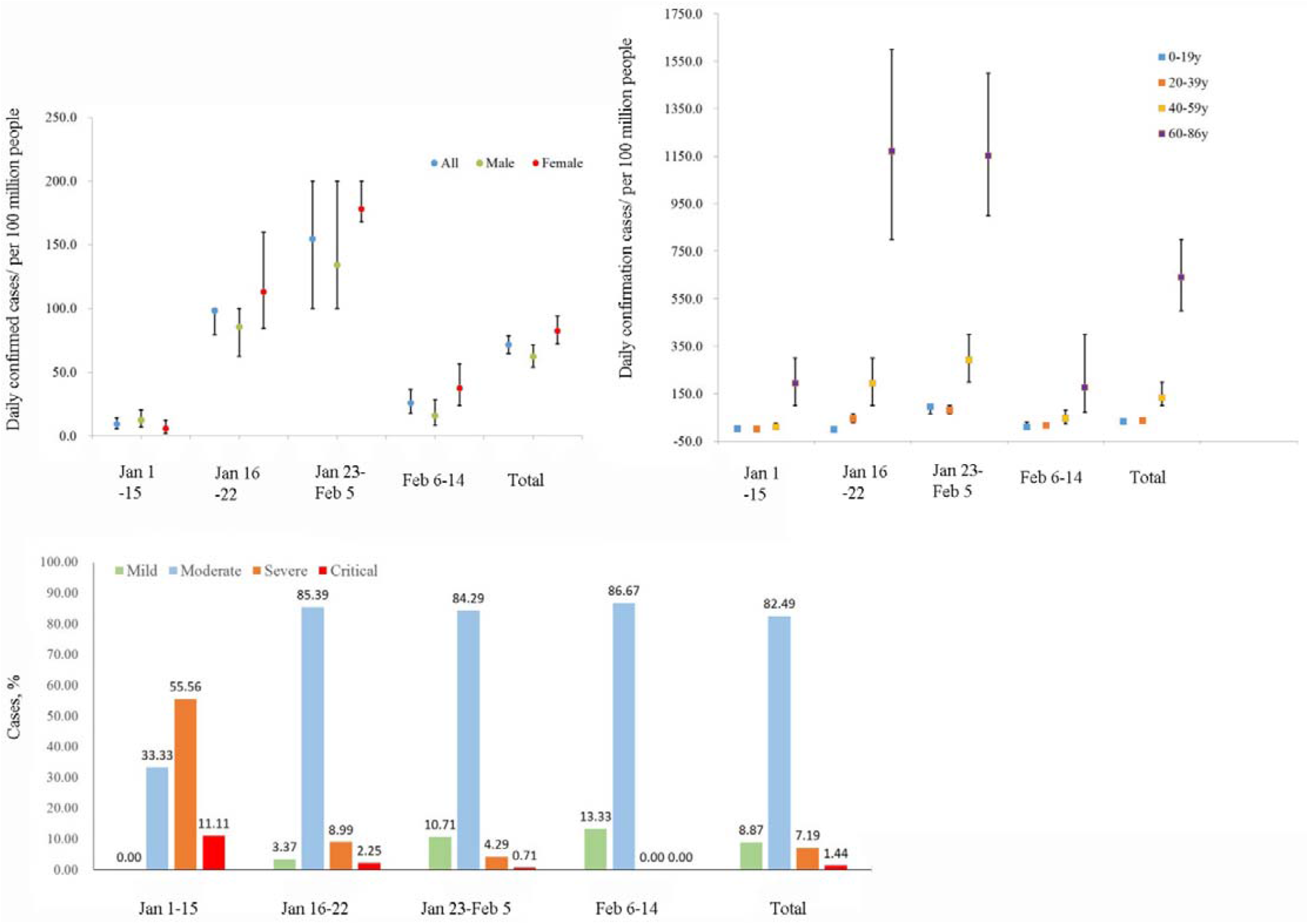
Daily confirmed cases in different groups by gender (A) and age (B). (C) The proportion of severity across the 4 time periods in Shenzhen, China. Error bars indicate 95% CIs.

**Table 1.** General characteristics of the confirmed COVID-19 cases in Shenzhen till February 14 by illness onset date

| Characteristics | January 1-15 | January 16-22 | January 23-February 5 | February 6-14 | Total |
| --- | --- | --- | --- | --- | --- |
| Total no. | 18 | 89 | 280 | 30 | 417 |
| Infection source-no. (%) |  |  |  |  |  |
| Input | 16(88.9) | 83(93.3) | 225(80.4) | 18(60) | 342(82.0) |
| Local | 2(11.1) | 6(6.7) | 55(19.6) | 12(40) | 75(18.0) |
| Exposure sources-no. (%) |  |  |  |  |  |
| Wuhan | 14(77.8) | 75(84.3) | 131(46.8) | 4(13.3) | 224(53.7) |
| Wuhan residents | 8(44.4) | 38(42.7) | 52(18.6) | 1(3.30) | 99(23.7) |
| Wuhan travelers | 6(33.3) | 37(41.6) | 79(28.2) | 3(10.0) | 125(30.0) |
| Hubei other than Wuhan | 1(5.6) | 6(6.7) | 64(22.9) | 9(30.0) | 80(19.2) |
| Shenzhen | 2(11.1) | 6(6.7) | 55(19.6) | 12(40.0) | 75(18.0) |
| Others | 1(5.6) | 2(2.2) | 30(10.7) | 5(16.7) | 38(9.10) |
| Identification model- no. (%) |  |  |  |  |  |
| Self-identified | 17(94.4) | 80(89.9) | 182(65.0) | 8(26.7) | 287(68.8) |
| Surveillance-identified | 1(5.6) | 9(10.1) | 98(35.0) | 22(73.3) | 130(31.2) |
| Sex-no. (%) |  |  |  |  |  |
| Male | 13(72.2) | 42(47.2) | 132(47.1) | 10(33.3) | 197(47.2) |
| Female | 5(27.8) | 47(52.8) | 148(52.9) | 20(66.7) | 220(52.8) |
| Median age(IQR)-year | 62.5(49.8-65.3) | 52(38-62.5) | 45(33-58) | 40(27.5-54.8) | 47(34-60) |
| Age group-no. (%) |  |  |  |  |  |
| 0-19 | 1(5.6) | 0(0) | 30(10.7) | 2(6.7) | 33(7.9) |
| 20-39 | 2(11.1) | 25(28.1) | 90(32.1) | 12(40.0) | 129(30.9) |
| 40-59 | 4(22.2) | 33(37.1) | 99(35.4) | 10(33.3) | 146(35.0) |
| 60-86 | 11(61.1) | 31(34.8) | 61(21.8) | 6(20.0) | 109(26.1) |
| Coexisting disorders-no.<br>(%) * | 2(11.1) | 23(25.8) | 65(23.2) | 6(20.0) | 96(23.0) |
| Cardiovascular diseases | 2(11.1) | 12(13.5) | 34(12.1) | 2(6.7) | 50(12.0) |
| Median incubation period<br>(IQR)-days ** | - | - | - | - | 5(2.5-9) |
| Duration from illness<br>onset to first medical visit<br>(IQR)-days† | 5(2-10) | 2(1-4) | 1(1-3) | 1(1-2) | 2(1-4) |
| Duration from illness<br>onset to hospitalization<br>(IQR)-days† | 10(7-12) | 5(3-8) | 4(2-6) | 1(1-4) | 4(2-7) |
| Duration from illness<br>onset to laboratory<br>confirmation (IQR)-days | 12(7-14 ) | 7.5 (4-10) | 4(3-7) | 1(1-6) | 5(3-8) |

†
\*Coexisting disorders include cardiovascular diseases, diabetes, lung diseases, immune-deficiency diseases, chronic hepatopathy and nephropathy.
\*\* The median incubation period was calculated in 47 local cases with clear exposure history.
† The time durations were calculated on the basis of 304 patients with symptom at onset, and the close contacts were excluded.

Among the total patients, about 197 (47.2%) were men and 220 (52.8%) were women (**Table 1**). The average daily confirmed cases per 100 million people (attack rate) drastically increased from 9.3 (95% CI 5.7 to 14.3) between January 1-15, to 98.2 (95% CI 79.3 to 100) between January 16-22, and to 154.4 (95% CI 100 to 200) between January 23-February 5, while declined to 25.7 (95% CI 17.7 to 36.2) after February 6 (**Fig 2A**). Similar trends were observed for men and women, with slightly higher attack rate for women during period 2, 3 and 4 and in the total cases. The exact values of the attack rates for each panel are shown in **Table S1**.

The median age of the patients was 47 years, with the majority (n=368, 88.2 %) of the cases aged 20 to 70 years (**Table 1**). A total of 33 (7.9%) patients were children under 20 years old, and the proportion reached 10.7% in period 3. The proportion of cases aged between 60~86 years decreased across time, while it showed an increasing trend for cases aged of 20~39 years. The attack rate increased with age, and the highest attack rate appeared in cases aged 60 to 86 years (640.44, 95% CI 500 to 800), followed by the cases aged 40 to 59 years (133.89, 95% CI 100 to 200) in any of the time period. For all age groups, the attack rate peaked in the third period and then decreased in the fourth period (**Fig 2B**). The exact values of the attack rates for each panel are shown in **Table S1**.

The clinical severity of the cases was classified into mild (n=37, 8.9%), moderate (n=344, 82.5%), severe (n=30, 7.2%) and critical (n=6, 1.4%) as shown in **Fig 2C**. The proportion of severe (severe/critical) cases decreased gradually across time, accounting for 66.7%, 11.2%, 5.0% and 0% in the four periods, respectively, while the proportion of mild/moderate cases increased over time. Logistic regression analysis was conducted to figure out risk factors for the severity of COVID-19. As shown in **Table 2**, results suggested that males were associated with a higher risk of severity (OR=2.11, 95% CI 1.04 to 4.28). Since there was only one severe case in the patients aged between 1~39 years, so we just compared the disease risk between the patients aged 40~59 and aged 60~86 years, and the results suggested that those older than 60 were more likely to be severe/critical (OR=2.99, 95% CI 1.41 to 6.31). Among the overall cases, 96 (23.0%) had at least one coexisting illness such as cardiovascular diseases, chronic pulmonary diseases or diabetes etc., and 50 (12.0%) had the cardiovascular disease history (**Table 1**). In the context of that, we found that the patients with co-existing disorders showed an increased risk for COVID-19 (OR=2.85, 95% CI 1.20 to 6.77). Specifically, patients with cardiovascular diseases showed a higher risk of COVID-19 (OR=2.31, 95% CI 0.99 to 5.39).

**Table 2.**
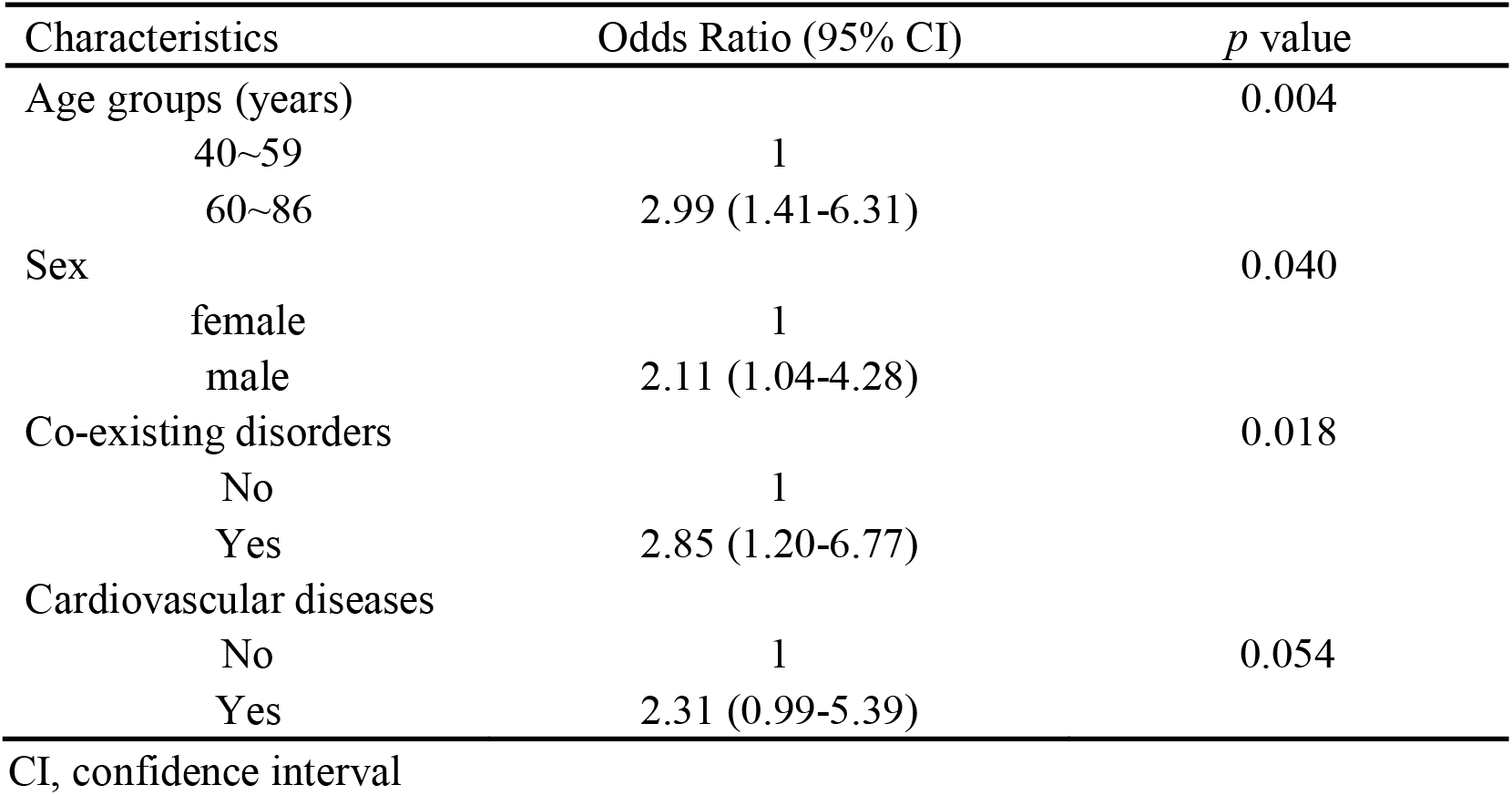
The associations of age, sex and co-existing disorders with the severity of COVID-19.

The residential distributions of the cases were shown in **Fig S2**. There are ten administrative districts in Shenzhen. The proportions of case number in Nanshan, Futian, Longgang, Bao’an, Luohu and Longhua districts accounted for 20.9%, 19.9%, 17.7%, 14.6%, 8.2% and 7.7% of the total cases, respectively. The proportion of cases in Nanshan decreased across time and dropped dramatically at period 4, while it showed an increased trend for Bao’an district. The attack rates for each district were shown in **Fig 3**. The epidemic started in the urban districts of Shenzhen and then gradually spread to the suburban districts across time.

**Figure 3.**
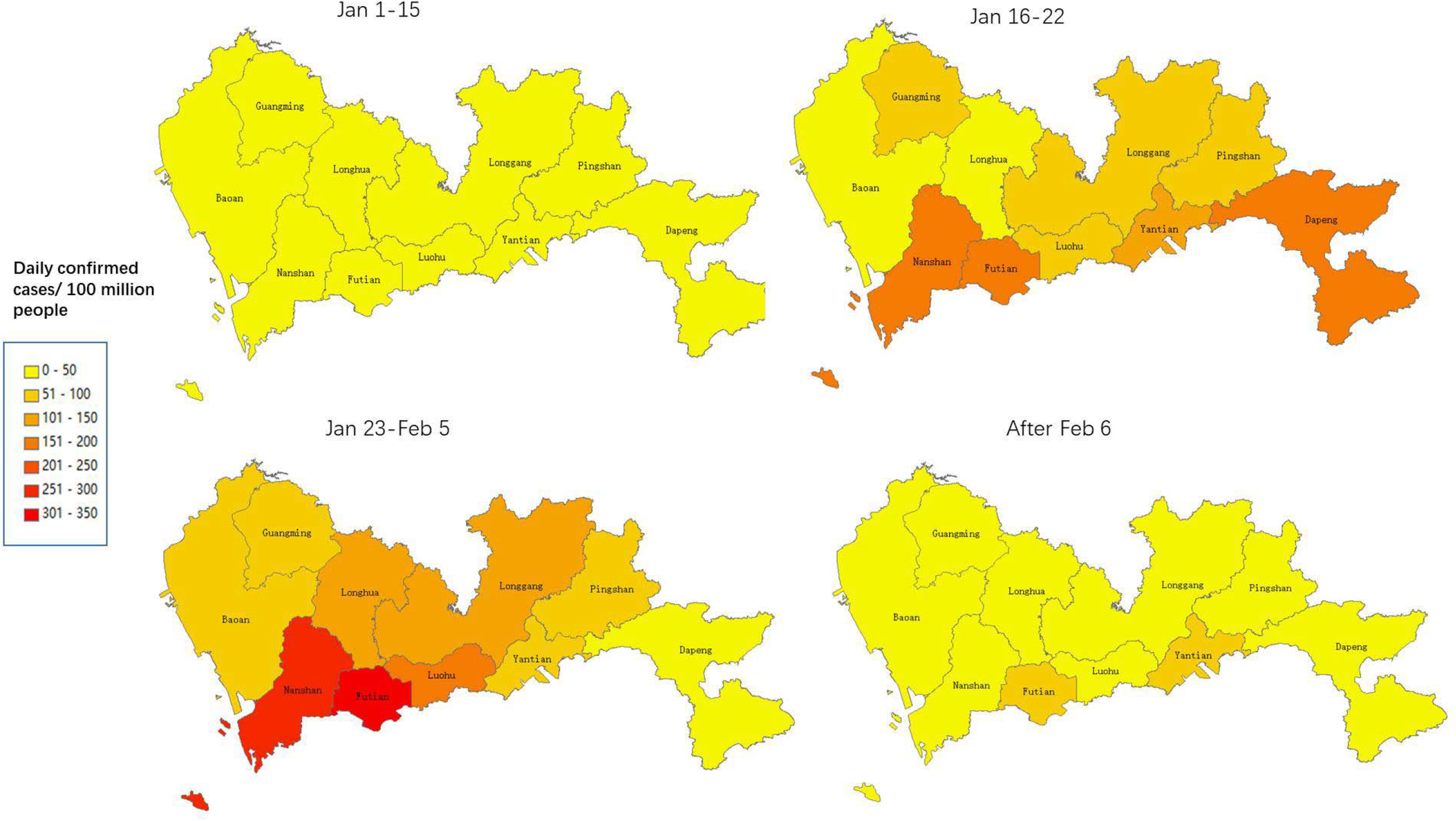
The geographic distribution of daily attack rates of COVID-19 across 4 time periods in Shenzhen, China. The daily rate of cases was indicated by the number of laboratory-confirmed cases per day per 100 million people, classified by each of the 10 districts in Shenzhen.

Using the detailed information of exposure time and illness onset of 47 local cases, the median of the incubation period of patients was estimated to be 5 days (IQR, 2.5 to 9), with the longest up to 17 days. Based on 304 cases with symptom at illness onset, we estimated the median time from illness onset to first medical visit, hospitalization and laboratory confirmation to be 2, 4 and 5 days for the overall patients, respectively (**Table 1**). The time intervals were all substantial during period 1, while the lag decreased across time, and it only took one day during period 4 for patients from illness onset to hospitalization or laboratory confirmation.

### Evaluating the effective of interventions in Shenzhen

The SEIAR model (**Fig S3**) was applied to evaluate the effect of interventions. The settings for parameters are described in **Table S2**. Among the input and local cases, about 40 and 14 patients were asymptomatic at illness onset, respectively. After excluding the asymptomatic patients, the SEIAR model in this study fitted observed data well *(χ*^2^ = 22.807, *P* = 0.993), as shown in **Fig 4A/B**. The transmission rate was 2.28 × 10^-8^ before January 23, and it decreased to 1.21 × 10^-9^ in the second period (**Table S2**). Accordingly, the *R_eff_* was 1.44 and 0.05 for the two periods, respectively, indicating a 96.5% decrease of *R_eff_* after the highest level emergency response was implemented in Shenzhen, Guangdong (**Fig 4A/B)**.

**Figure 4.**
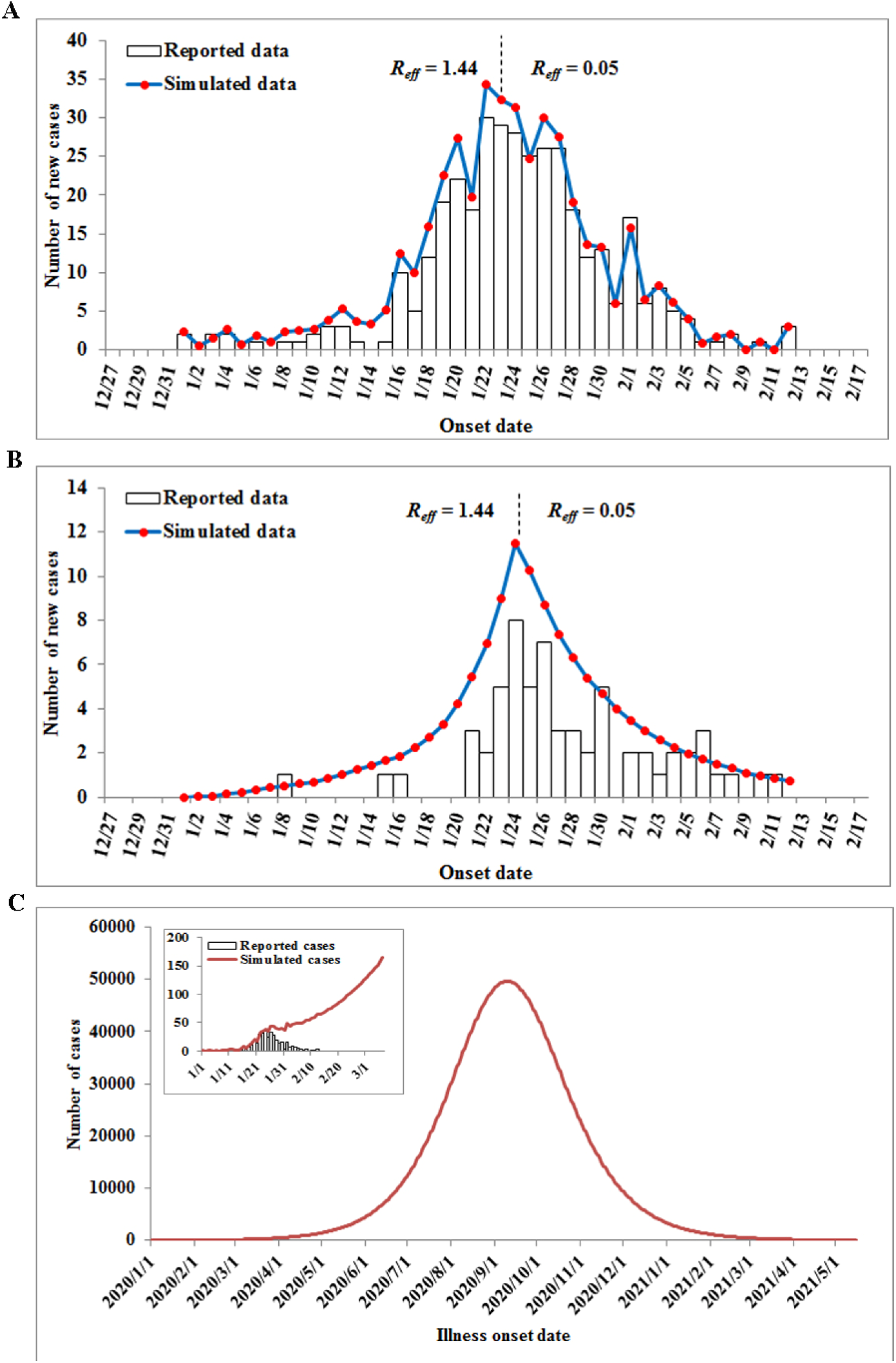
Comparison between simulated results by the SEIAR model and the reported data (including imported and local cases) of COVID-19 in Shenzhen, China. Modeling fitting in total cases (A) and the local cases (B); (C) Prediction of the epidemic of COVID-19 using the parameters from the first period (January 1-22).

The cumulated number of symptomatic infections was estimated to be 2671 till February 29, if the transmission rate of the first period continued (**Fig 4C**), indicating that the highest level emergency response of Shenzhen might lead to 86.3% decrease of the total symptomatic patients (2671 vs. 366). We also estimated the cumulated number of local symptomatic infections to be 2367 till February 29 if no intervention was implemented on January 23, inferring a decrease of 97.4% of the local symptomatic patients (2367 vs. 62). Without intervention, the transmission would reach a peak on September 9, 2020 and go to the end on July 1, 2021 with an attack rate of 45.25% for the overall population.

We also simulated four additional scenarios (*R_eff_* = 1.80, 2.20, 2.60, and 3.00) of the transmission of COVID-19 in Shenzhen City, China. Our modeling results showed that if the *R_eff_* was set to be 1.80 in the first time period, the transmission would reach a peak on June 21, 2020 and go to an end on February 12, 2021 in condition of no highest level emergency response was initiated, with a total attack rate of 60.81%. If we set *R_eff_* = 2.20, the transmission would reach a peak on May 12, 2020 and go to an end on November 7, 2020 with attack rate of 70.04%. If we set *R_eff_* = 2.60, the transmission would reach a peak on April 20, 2020 and go to an end on October 4, 2020 with attack rate of 75.11%. If we set *R_eff_* = 3.00, the transmission would reach a peak on April 6, 2020 and go to an end on August 18, 2020 with attack rate of 78.07% (**Fig S4**).

## Discussion

In this study, we depicted a comprehensive picture of the general characteristics of 417 laboratory-confirmed COVID-19 cases in Shenzhen till February 29. Based on the observed data, we further conducted a modeling study to evaluate the effect of control measures on the transmission of COVID-19. Here, we show that the highest provincial emergency response in Guangdong combined with the travel restrictions in Wuhan have effectively mitigated the spread of SARS-CoV-2 in Shenzhen, China. Our findings might provide important parameters for further analysis in situation of pandemic transmission of COVID-19, including predicting the future spread of the virus worldwide, and evaluating the effect of non-pharmaceutical interventions in other highly-affected countries.

Most cases in our study were middle-aged, and the median age was 47 years old, which is consistent with a previous report regarding 1099 confirmed patients extracted from 552 hospitals in 30 provinces (Guan et al., 2020). Different from the previous findings in Wuhan (Li et al., 2020a), we found the proportion of young patients (20~39 years old) was slightly higher than the elderly (60~86 years), probably because that the average age of people in Shenzhen is relatively younger compared with other cities. The attack rate kept increasing before February 6 and declined afterwards for all age groups. Thirty-three children under 20 years old were infected, and we observed that most of them (94%, 31/33) were affected in the clustering, as warned by WHO report that the infection risk would increase by clustering when the family members were required to stay at home (Report of the WHO-China Joint Mission on Coronavirus Disease 2019). The situation was worse in Wuhan, where the daily confirmed cases per million people kept increasing over time for children less than 20 years, and the infants less than one year old showed the highest attack rate among the children (Pan et al., 2020). However, due to the limited sample size, we only compared the attack rates between children of 0~5 and 6~19 years. The children aged 0~5 years showed a higher attack rate compared with those of 6~19 years old, probably because younger children have lower immunity (Wei et al., 2020). The attack rate of the elderly was the highest among all age groups, and they were associated with increased risk of severity of COVID-19. Taken together, specific attention should be paid to the vulnerable population including younger children and the elderly people, in order to restrain the transmission of the virus.

The first case of unexplained pneumonia was identified in Shenzhen on January 8. The epidemiological investigation suggested a travel history to Wuhan but without exposure history to Hua-Nan seafood market, the original source of the virus. Meanwhile, one of the family members who had never been to Wuhan also got illness onset. Therefore, this might provide critical evidence for the human to human transmission conclusion before the official announcement on January 20 by China government (Chan et al., 2020). The emergency response measures of the disease control system were activated promptly by Shenzhen CDC on January 10, and the joint prevention and control system for serious infectious diseases was started on January 16. According to the epidemic curve of Shenzhen, since January 17, 2020, the case number climbed with time, and it reached a plateau during January 22~27, then gradually decreased with oscillating afterwards, indicating that the large population migration during Chinese Lunar New year has facilitated the spread of virus outside Wuhan. Before the travel restriction on January 23, the input cases accounted for 92.5% of the total patients in the first period, mainly from Wuhan city (83.2%), and only 8 (7.5%) local patients were reported. After January 23, the imported cases from Wuhan dropped to 43.5%, while the proportion of local cases increased to 21.6%, indicating the spreading of virus in local. The Guangdong province initiated the highest emergency response on the same day with Wuhan *cordon sanitaire* policy. A recent study reported that the considerable countermeasures implemented in China substantially mitigated the spread of COVID-19 (Kraemer et al., 2020). Here, we applied the SEIAR model to estimate the effect of control measures in Shenzhen on the epidemic of COVID-19. The parameters including CFR, proportion of asymptomatic patients, incubation period and duration from illness onset to hospitalization were all calculated using the field epidemiology investigation data.

It has been commonly recognized that SARS-CoV-2 may have a lower CFR than SARS-CoV or Middle East respiratory syndrome–related coronavirus (MERS-CoV). We found a much lower CFR of 0.72% than reported recently (Guan et al., 2020; Chen et al., 2020a; Huang et al., 2020). However, considering the fact that there were substantial patients who did not seek medical care due to mild illness or shortage of medical resources in Wuhan, which might lead to an overestimation of CFR, we supposed that the CFR in Shenzhen (<1%) was closer to the reality. The early isolation, diagnosis and clinical management of the patients might also have greatly contributed to the low mortality in Shenzhen.

Accumulating evidence have shown that the asymptomatic patients can also be infectious for susceptible population (Bai et al., 2020), and a large proportion of the total infections might be mediated through the un-documented infections in China (Li et al., 2020b). Therefore, figuring out the proportion and the transmission rate of asymptomatic cases is critical for the prevention and control of COVID-19 worldwide. The proportion of asymptomatic cases was estimated to be 34.6% onboard the Princess Cruise ship (Mizumoto et al., 2020). What is more shocking is that researchers used the reported data in China in conjunction with mobility data and inferred that appropriately 86% of all infections in China were undocumented before the travel restrictions since January 23 (Li et al., 2020b). As scientists were setting out on estimating the proportion of people with mild or no symptoms (Qiu, 2020), we estimated the ratio of asymptomatic cases to be about 17% in Shenzhen, by calculating the proportion of all asymptomatic cases among the clustering cases using the field epidemiological investigation data. The ratio is much lower than the previous studies, probably because the cases with mild symptoms were not included for calculating; second, since the medical resources were relatively sufficient in Shenzhen, the medical care-seeking behaviors were more active in the infected people. Moreover, the early public health response and early isolation of asymptomatic patients by Shenzhen government also restrained the spread of the virus. Nevertheless, the proportion of asymptomatic infections needs to be confirmed in the future serologic studies as soon as possible.

The transmissibility of people with mild or no symptoms is another important question that scientists are rushing to solve (Qiu, 2020). A German-based research showed that some COVID-19 patients had very high levels of virus in throat swabs while their symptoms were mild (Woelfel, 2020). Another study of China also detected high viral loads in the cases without symptoms, who shed a similar amount of virus to those with symptoms (Zou et al., 2020). Shen et al. evaluated 24 asymptomatic patients in Nanjing, China and they found that the communicable period could be as long as three weeks, and the people infected by those asymptomatic could develop severe illness (Hu et al., 2020). These findings suggested that some infected people can be highly communicable when they have mild or no symptom. A recent study constructed a networked dynamic meta-population model and Bayesian inference using the data of reported infection in China and the mobility data. They found that the transmission rate of undocumented infections was 55% of the reported infections (Li et al., 2020b). In the context of that, according to the previous studies, here, in order to simulate the cases in condition of no intervention was implemented on January 23, the transmission rate of asymptomatic patients was set to be 50% of the symptomatic patients conservatively, as we previously reported in a modeling study to simulate the phase-based transmissibility of SARS-CoV-2 (Chen et al., 2020b).

The median incubation period was 5 days, and in most cases it ranged from 1~14 days, except for one case whose incubation period was about 17 days. Consistently, one previous study also demonstrated that about 10% of patients with COVID-19 would not develop symptoms until more than 14 days after infection (Jing et al., 2020), inferring that the general population should still be cautious of self-protection even after 14 days’ quarantine. Hospitalization is critical for disease control, and we observed that the delays between illness onset and first medical visit, hospitalization or diagnosis confirmation all decreased across time, indicating the effectiveness of control measures by Shenzhen government. Increased knowledge of the virus and extensive news coverage among the general population might have already facilitated the seeking behaviors of medical care. In addition, the supply of medical sources, including the viral identification assays, as well as the more active surveillance of close contacts and surveillance at the traffic barriers, further increased the capacity of suspected population identification and disease confirmation. In the context of that, as we observed in the investigation data, the proportion of cases identified by surveillance also showed an increasing trend across time.

Amounting evidence have reported varied basic reproduction number (*R*_0_) (range 1.40 to 6.49,) for COVID-19 due to different datasets, time periods or the statistical methods (Li et al., 2020a; Liu et al., 2020), and it has reached a consensus that the transmissibility of COVID-19 is similar or higher than that of SARS-CoV in 2003, with *R*_0_ estimated to be around 2.2 to 3.6 (Lipsitch et al., 2003). Our estimation of the effective reproduction number *R_eff_* =1.44 in the first period (before January 23) was even lower; however, it might not reflect the basic reproduction number, because several important interventions have been conducted by then. Since the beginning of January 2020, Shenzhen CDC has initiated the emergency response for unexplained pneumonia, and the local government implemented the joint defense and control mechanism after January 16, 2020, which might affect the transmissibility of the virus. In the context of that, the parameter *R_eff_* but not *R*_0_ was adopted for transmissibility assessment of the first period in Shenzhen. On January 23, the first-level provincial emergency response was implemented in Guangdong combined with the policy of cordon sanitaire in Wuhan city. The control measures of the provincial emergency responses include strengthening the joint prevention and control mechanism; banning public gathering; effective management of ascertained cases, suspected cases and close contacts; implementing quarantine on transportation; strengthening medical treatment mechanism; protecting the medical and health workers; focusing on the key populations, especially the elderly, students, faculties and return employees; strengthening the health education and news coverage to the general population, etc. To control the spread of the virus in communities, the “five 100%, ten must “policy was executed in Shenzhen since February 6, with a model of community workstations, polices and physicians combined together. Noticeably, the *R_eff_* dramatically decreased from 1.44 to 0.05 in the second time period, suggesting that the comprehensive measures implemented in Shenzhen was clearly successful in mitigating the spread and reducing local transmission.

Some limitations of this study need to be addressed. First, the proportion of asymptomatic patients might have been overestimated, since part of the patients might develop symptoms after they were confirmed as COVID-19 cases. Therefore, the serologic studies are warranted to confirm our estimates. Second, the transmissibility of people with no symptom was not clear till now; we set the parameter of transmission rate to be half of the symptomatic patients based on limited previous reports, which might reduce the accuracy of our results. Third, the control measures were evaluated as a whole in this study, and we could not determine the impact of each intervention individually.

In summary, the intensive non-pharmaceutical control measures have been implemented to restrain the spread of COVID-19 in Shenzhen, including the early emergency response of Shenzhen authorities and CDCs, early application of designated hospital and centralized isolation places for suspected cases and close contacts, the highest level emergency response in Guangdong, and the strict control measures for community transmission, etc. The epidemic curve and our modeling estimates suggested that the containment efforts implemented since the beginning of January, 2020 were clearly effective in mitigating the transmission of COVID-19, especially for the provincial first-level emergency response of Guangdong. These were encouraging for the combat against global COVID-19 outbreak at the time when no effective drug or vaccine for this new infectious disease has been developed. Besides, much further work is warranted by government to balance the public health of disease control and the negative effects on economy and society at large.

## Materials and Methods

### Source of data

The general information of COVID-19 cases were extracted from the field epidemiological survey reports and the National Infectious Disease Information System between January 1, 2020 and February 29, 2020, including age, sex, residential district, co-existing disorders, illness severity, source of infection, date of illness onset, hospitalization, and laboratory confirmation in bio-samples.

### Study definition

Cases were diagnosed and classified as mild, moderate, severe, and critical according to the sixth editions of the Diagnosis and Treatment Scheme for COVID-19 released by the National Health Commission (NHC) of China (National Health Commission of the Peoples’ Republic of China). A case was ascertained as COVID-19 if he/she was tested positive for SARS-CoV-2 virus, either by real-time reverse transcription polymerase chain reaction (RT-PCR) assay or by high-throughput sequencing in the nasal/pharyngeal swab specimens. Only laboratory confirmed cases were included in this study. The cases were identified by medical care seeking or by surveillance. The former referred to those who sought medical care at illness onset, and the latter cases referred to those identified by surveillance, including close contacts and recent travelers from Hubei; fever monitoring at the traffic barriers, and people with symptoms reported by community (Wen et al., 2020). The incubation period was defined as the time interval between the average date of contact with potential infection source and the date of symptom onset. The incubation periods were calculated based on 47 local patients who had clear information about the exact date of exposure and symptom onset. The time intervals between illness onset and first medical visit, hospitalization and laboratory confirmation of the cases were estimated on the basis of 304 cases with symptom at onset, and the close contacts were excluded in the analysis.

### Definition of different time periods

To better illustrate the full spectrum of transmission dynamics for COVID-19, we divided the epidemic into several periods based on the illness onset day and the non-pharmaceutical interventions that might affect virus transmission, as demonstrated in **Fig 1**. Since the NHC of China acknowledged a novel coronavirus as the cause of unexplained pneumonia in Wuhan, the Shenzhen authorities have implemented counter-measures since January 5, 2020, and the CDCs have activated the emergency response since January 10, 2020. In addition, the Shenzhen government has strengthened the management of fever clinics, and designated the hospital for treatment since January 13, 2020. Moreover, the virus detection kits were available in Shenzhen on the same day. The second period refers to *Chunyun* of January 16 ~22, 2020, when a large population migrated for the Chinese New Year and might have speeded the spreading of COVID-19. Several important interventions were carried out during this period, including starting the joint defense and control mechanism for major infectious diseases on January 16, as well as applying the concentrated quarantine point for suspected cases and close contacts on January 21. During the third period after January 23, besides the Wuhan *cordon sanitaire* policy which aimed to restrain population migration, the Guangdong government has activated the highest-level emergency response on January 23, followed by a series of rigorous measures, including comprehensive community screening and strict access management; curbing gathering activities and social distancing; compulsory mask-wearing in public places; temperature monitoring in traffic barriers; effective isolation for suspected cases, close contacts and treatment for ascertained cases; protecting medical and health workers from infection and shutting down schools and training institutions, etc. Since February 6, the “five 100%, ten must” policy was implemented aiming to control the community spreading of COVID-19, indicating a turning point of the control strategy switching from input prevention to community transmission restraining in Shenzhen. The detailed information about the highest level emergency response of Guangdong and the “five 100%, ten must” policy were described in the **Supplementary material**.

In summary, the COVID-19 outbreak in Shenzhen was divided into four periods (January 1-15, January 16-22, January 23-February 5, and February 6-14, respectively).

### Modeling

The SEIAR model (**Fig S3**) was applied to simulate epidemic data and evaluate the effect of intervention measures, accounting for a substantial proportion of asymptomatic patients. Since the case numbers in period 1 and 4 were small according to the classification results, we combined period 1 and 2 together as the first period for modeling analysis, and the period 3 and 4 were combined as the second period. We classified the population into five compartments, including susceptible individuals (S), latent cases (E), reported infections (I), asymptomatic cases (A) and removed individuals (R). The dynamics of five compartments and the key parameters of the model across time were demonstrated by ordinary differential equations:

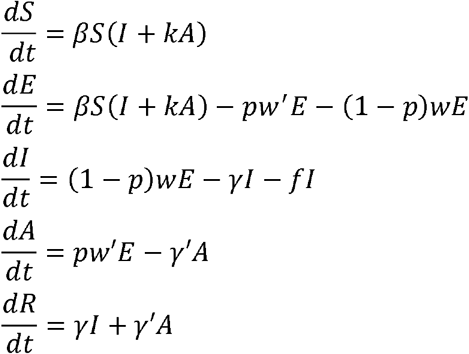

In the equations, *β* was the transmission rate; *k* was the ratio of transmission rate of asymptomatic cases compared with symptomatic cases; *p* was the proportion of asymptomatic cases; *ω* and *ω′* were the reciprocal of incubation period and latent period, respectively; *r* was the removal rate of ascertained cases, which is the reciprocal of the time interval from illness onset to hospitalization; *r’* was the removal rate of the asymptomatic cases, which was set to be the same with symptomatic patients; *f* was the case fatality rate (CFR). The effective reproductive number *R_eff_*, was computed for each period. The *R_eff_* was calculated as:

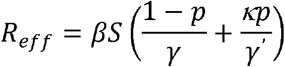

The proportion of asymptomatic infections, incubation period, CFR and the time duration from illness onset to hospitalization were calculated by the field epidemiology investigation data. We set latent period and incubation period as 5 days, so the parameters *ω* and *ω′* were both 0.2. The durations from illness onset to hospital isolation were 6 and 4 days for the two time periods, respectively. The removal rate of asymptomatic cases was set to be the same with that of symptomatic patients according to one of our previous study (Chen et al., 2020b), so the parameters *r/r’*was 0.17 and 0.25 for the two periods, respectively. As we previously reported, two hundred and forty-seven of the 417 patients (59.2%) were identified in 92 clusters (Wen et al., 2020).The proportion of asymptomatic infections (*p*) was set according to the proportion of asymptomatic patients among clustering cases, which is 0.17 (42/247). The CFR was set to be 0.00718 (3/417). We set the transmission ratio between asymptomatic and symptomatic cases to be 0.5, assuming half of the transmissibility for asymptomatic patients compared with symptomatic cases, according to one of our previous study (Chen et al., 2020b). The population of Shenzhen was set according to Shenzhen Statistical Yearbook 2018.

### Statistical analysis

The COVID-19 cases with onset time ranged from January 1, 2020 to January 14, 2020 were included in this study. Continuous variables were denoted as median (interquartile range), and categorical variables were denoted as frequency. The attack rate was defined as the number of infections per day per 100 million people, analyzed by sex, age and residential districts across time periods. The calculations used the number of cases in each period divided by the days in each period (18, 89, 280 and 30 days, respectively) and the subtotal population size referring to the Shenzhen Statistical Yearbook 2018. The geographical distributions of daily rates of COVID-19 cases across Shenzhen in the 4 periods were depicted by ArcGIS software version 10.6 (Environmental Systems Research Institute Inc). We evaluated the association of age, sex and co-existing disorders with disease severity (mild/moderate versus severe/critical). Odds ratios (ORs) and 95% confidence intervals (CIs) were calculated.

The imported cases were simulated as transmission sources and the secondary cases were applied for the curve fitting. The procedures of the simulation and curve fitting were performed by Berkeley Madonna 8.3.18 (developed by Robert Macey and George Oster of the University of California at Berkeley. Copyright ©1993-2001 Robert I. Macey & George F. Oster, CA, USA). The simulation methods (Runge-Kutta method of order four with tolerance set at 0.001) were described in the previous reports (Chen et al., 2019; Chen et al., 2014; Zhang et al., 2020). The goodness of fitting was assessed by Chi-square (*χ*^2^) value using SPSS 21.0 (IBM Corp, Armonk, NY, USA).

## Compliance and ethics

The authors declared that they have no conflict of interest. This study was conformed with the Helsinki Declaration of 1975 (as revised in 2008) concerning Human Rights, and was approved by the ethics committee of the Shenzhen Center for Disease Control and Prevention. The informed consent for collection of epidemiological data from COVID-19 patients was granted by the NHC of China as part of the infectious disease field epidemiology investigation. All personal information was removed for privacy protection.

## Acknowledgements

The authors would like to thank the patients and all the staff from Shenzhen CDC for data collection. This work was supported by the Open Fund of Shenzhen Bay laboratory (SZBL202002271004) and SanMing Project of Medicine in Shenzhen (SZSM201811070).

